# Strain-level characterization of health-associated bacterial consortia that colonize the human gut during infancy

**DOI:** 10.1101/2023.12.16.23300077

**Authors:** Samuel S. Minot, Koshlan Mayer-Blackwell, Andrew Fiore-Gartland, Andrew Johnson, Steven Self, Parveen Bhatti, Lena Yao, Lili Liu, Xin Sun, Yi Jinfa, James Kublin

## Abstract

**Background:** The human gut microbiome develops rapidly during infancy, a key window of development coinciding with maturation of the adaptive immune system. However, little is known of the microbiome growth dynamics over the first few months of life and whether there are any generalizable patterns across human populations. We performed metagenomic sequencing on stool samples (n=94) from a cohort of infants (n=15) at monthly intervals in the first six months of life, augmenting our dataset with seven published studies for a total of 4,441 metagenomes from 1,162 infants.

**Results:** Strain-level *de novo* analysis was used to identify 592 of the most abundant organisms in the infant gut microbiome. Previously unrecognized consortia were identified which exhibited highly correlated abundances across samples and were composed of diverse species spanning multiple genera. Analysis of a cohort of infants with cystic fibrosis identified one such novel consortium of diverse *Enterobacterales* which was positively correlated with weight gain. While all studies showed an increased community stability during the first year of life, microbial dynamics varied widely in the first few months of life, both by study and by individual.

**Conclusion:** By augmenting published metagenomic datasets with data from a newly established cohort we were able to identify novel groups of organisms that are correlated with measures of robust human development. We hypothesize that the presence of these groups may impact human health in aggregate in ways that individual species may not in isolation.

## Background

Early-life colonization of the human gut by microorganisms can have long-term implications for physiology and disease[1–3]. Species- and strain-level analyses suggest that most taxa can be inherited from the mother during vaginal birth, and microbial transfer is likely reduced in infants born by Caesarean delivery or by those treated with antibiotics[4–6]. Disruptions to natural bacterial exposures and microbiome development (e.g., by Caesarian section delivery, excessively sterile environment, or antibiotic-treatment) are associated with increased susceptibility to inflammatory and metabolic diseases, and intervention studies in animal models have defined key pre- and post-natal developmental windows during which the developing microbiome affects important immune processes, such as tolerance induction.

Key knowledge gaps remain concerning the immune phenotypes of at-risk infant populations and how early-life complications, such as microbiome disruption, malnutrition, and pathogen exposures, alter immune ontogeny and lead to vaccine response deficiencies in some children. Emerging evidence suggests that individual variation in response to infection or vaccination may be influenced by past viral and bacterial exposures, which shape the immune system and can establish pre-existing immune-reactivity[7–10].

Murine systems and longitudinal human birth cohorts have defined critical neonatal windows in which the intestinal microbiome stimulates immune maturation and provides colonization resistance to protect against infectious and immune-mediated disease[3, 11–28]. While neonatal taxa-immune pathways remain to be fully elucidated, the acquisition of Clostridiales taxa in early-life is clearly vital [2, 29–44]. Clostridiales provide colonization resistance[13], stimulate immune-regulatory responses[18, 26, 45–47], and activate IFN-mediated lung protection[48]. A failure to acquire Clostridiales taxa, especially *Ruminococcaceae, Lachnospricaeae* and Clostridium Cluster XIVa, represents the major deficiency of the CF infant microbiome, a finding that is highly reproducible across multiple independent cohorts, including the most extensively characterized BONUS cohort[30, 31, 35, 37, 44].

Longitudinal studies of birth cohorts – so far conducted predominantly in North America and Europe – have begun to characterize compositional changes to the gut microbiome that occur in the first years of life. These studies have relied primarily on amplification of the bacterial 16S ribosomal RNA gene or, more seldomly, whole genome sequencing[4, 6, 49–51]. These longitudinal studies, along with one major cross-sectional study[52], have demonstrated that there is considerable inter-individual and temporal variation in the neonatal and infant microbiome community starting from birth and extending to approximately three years of age. During this period the microbiome gains richness and stability to form a microbial community that is more reflective of the adult microbiome[4, 6, 49–51, 53]. This represents a general transition where bacteria specialized to the aerobic neonatal gut (e.g., *E. coli*) or for growth on complex sugars in breastmilk (e.g., Bifidobacterium and Veillonella) and are outcompeted by organisms found more commonly in the adult gut microbiome, such as Bacteroideceae and Rumminococcaceae[4, 6, 50]. This is reflected in the metagenomic composition of the bacterial communities, with genes involved in milk oligosaccharide metabolism giving way to those better suited to solid foods, such as fiber degradation[4, 6, 49].

While these studies have elucidated general trends in infant microbiome development, most prior studies are limited by low density of fecal sampling in the first 6 months of life when temporal intraindividual variation in the microbiome is highest and exposures to the immune system are particularly impactful. Furthermore, bioinformatic approaches have focused predominantly on identifying microbiome community states that are reflective of specific ages and which are generalizable across individuals. In contrast, these approaches offer more limited insight into the growth dynamics of individual taxa or clusters of interacting consortia. It is now evident that the path of individual microbiome development is highly variable across infants.

For example, a recent analysis of transitions between ten different microbiome community states in early life observed great diversity in the patterns of transitions between states. In fact, the most common transition pattern was only observed in 20% percent of infants, with the remaining 80% of infants displaying unique maturation transition patterns.[54]

Recognizing that the microbiome may not conform to consistent community states, an alternate ontological approach is to identify the subsets of microbial organisms which are found together in concert, as would be expected from a group of Bifidobacteria which jointly metabolize human milk oligosaccharides. From an informatic point of view, we approach the identification of microbial consortia by testing for organisms with correlated abundances across large numbers of microbiome samples[55]. When organisms are more likely to be found together than would be expected by random assortment, we may hypothesize that there is a shared underlying biological process which is jointly driving their growth and survival.

In human microbiome research, detection of well-studied bacterial species and genera can reveal considerable information about environmental conditions present during health and disease conditions of the host; however, there are limitations in taxonomic-centered approaches that orient analysis around finding associations with host characteristics and relative abundance of bacterial groups agglomerated by phylogenetic clade. Critically, aggregating organismal relative abundances within phylogenetic clades (i.e., summarizing microbiome features to the genus, family, or order level) becomes less informative as physiology and metabolism of bacteria within taxonomically derived grouping can vary greatly. However, strain-level analysis suffers from high-dimensionality and high sparsity of features between samples. Thus, a major challenge in analysis of microbiome is finding a flexible unit of analysis that permits detection of consistent and interpretable ecological changes in the host via phylogenetic-independent agglomeration of co-abundant organisms.

Thus, to gain a better understanding of dynamics of microbiome development in the critical development period between 1 to 6 months of age, we conducted such a gene-level microbiome analyses on stool samples collected monthly in a longitudinal mother-infant birth cohort[56]. Because the aggregate gene content of the gut microbiome is comprised of tens or hundreds of millions of genes[57], a meaningful embedding in lower dimensional space is helpful for comparisons across samples. For this purpose, we use Co-Abundant Gene Groups (CAGs)[58, 59] which represent sets of genes that are expected to be found together in the same genetic element (chromosome, plasmid, virus, etc.) across all of the samples in the collection. To increase the total amount of biological information used for CAG construction, we augmented the data from our own cohort with additional metagenomic data from seven published infant microbiome datasets[4, 6, 49–53], for a total of 4,441 biological samples in the combined dataset. We used a reproducible pipeline, *geneshot* [60], for constructing and quantifying CAGs to describe groups of organisms that colonize the gut according to patterns of correlated abundance which are reproduced across cohorts. We describe these groups of microbes which are present or absent in concert as previously-unrecognized microbial consortia, which may help researchers more succinctly describe the patterns of rapid turnover which are observed during the first six months of life. Moreover, parallel analysis of a published cross-sectional cohort[53] identified one such consortium whose presence was strongly correlated with infant growth rate. We propose that this dynamic growth variation may underlie altered immune development between individuals and associated susceptibility to immune-mediated disease in later life, and therefore that CAG-based analysis of microbial consortia would be a useful approach for the analysis of existing and future longitudinal birth cohorts.

## Results

### *De novo* metagenomic analysis identifies 592 bacterial genomospecies in the infant gut

To quantify the relative abundance of microorganisms present in the gut during early human life, metagenomic whole-genome shotgun sequencing (WGS) data were generated from the total DNA isolated from stool samples collected from cohort of healthy infants (n = 15) at monthly intervals from birth until 6 months. To mitigate the inherent stochasticity of metagenomic sequencing, we sought to increase generalizability of gene-level analysis by including other publicly available deeply sequenced microbiome samples from other infant cohorts. the most that are In all, we analyzed metagenomic data from the seven largest published infant microbiome datasets[4, 6, 49–53], for a total of 4,441 biological samples in the combined dataset (Fig. 1A, Table 1, Table S1, S2). Gene-level metagenomic analysis was performed by: (1) generating a gene catalog via *de novo* assembly and centroid clustering (based on 90% amino acid identity); (2) estimating the relative abundance of organisms encoding each individual gene via short-read alignment; and (3) grouping genes with correlated abundances into co-abundant gene groups (CAGs) via iterative greedy single linkage clustering[60]. Thus, the primary unit of measurement for downstream analyses was the relative abundance of each CAG in a particular sample, which is an estimate of the relative abundance of the organisms contained within that sample encoding the genes in that CAG. Because the groups of genes contained in each CAG have highly correlated abundances, they are predicted to be contained within the same genomic context with the metagenome, representing the complete or partial core genome of a set of closely related isolates or strains[58].

**Figure 1.**
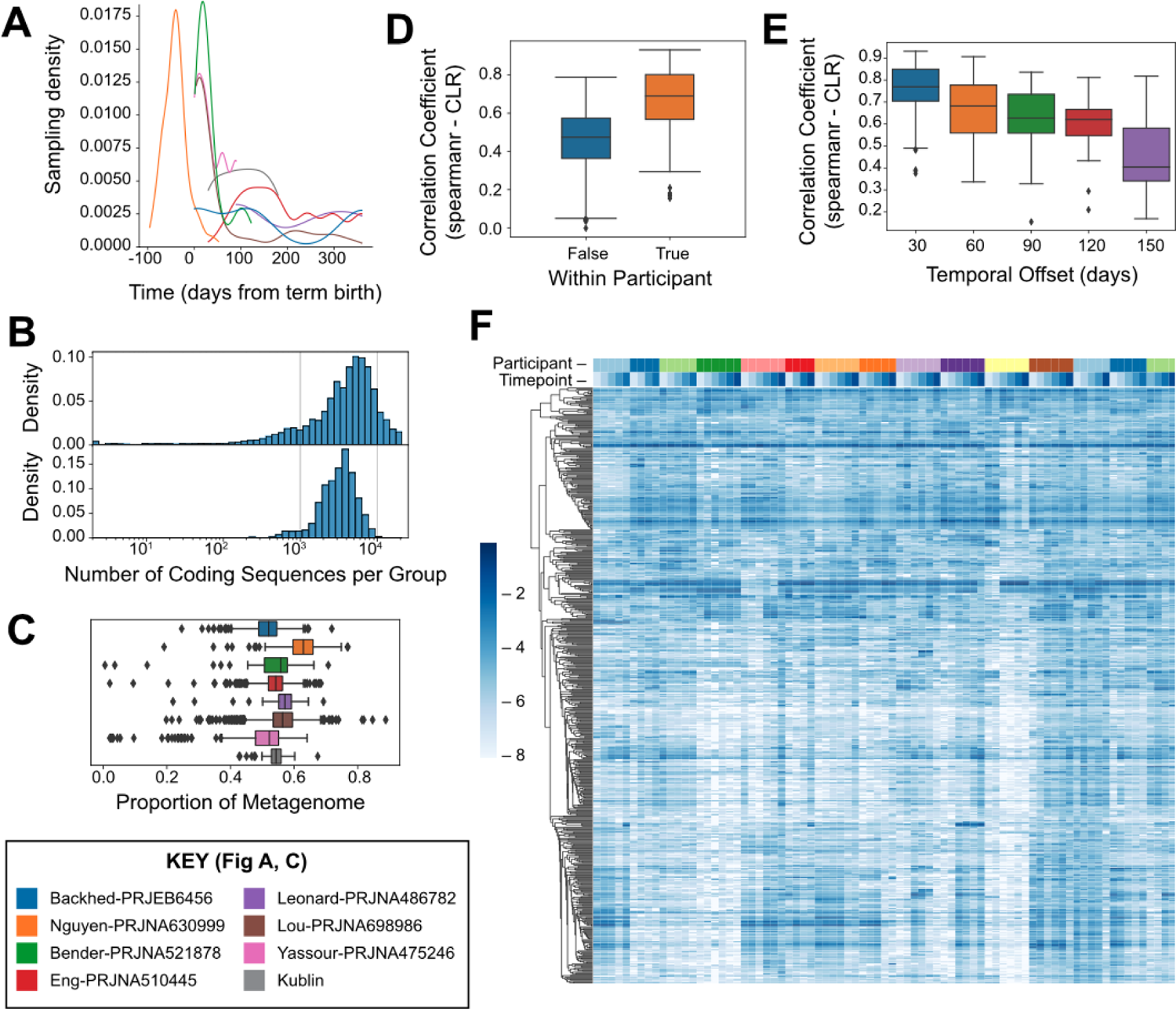
Quantification of 592 microbial genomospecies from 0-6 months across 8 studies. A) Density of sample collection per study over time relative to term birth (40 weeks of gestational age). B) Distribution of genomospecies genome sizes (number of coding sequences, top) in comparison to NCBI prokaryotic reference genomes (bottom). C) Proportion of metagenomic sequence data from each sample which can be unambiguously assigned to any one of the 592 bacterial genomospecies, compared across studies. D) Within-versus intra-participant variation in metagenome similarity estimated using the Spearman correlation coefficient of Centered-Log Ratio (CLR) abundances. E) Comparison of within-participant variation in metagenome similarity between samples at varying time intervals estimated using the Spearman correlation coefficient of CLR abundances. F) Comparison of microbiome composition within the samples collected for this study, with participants (n=15) and timepoints (n=6) indicated on the top marginal axis. Dendrogram indicates hierarchical clustering of the 592 genomospecies based on similarity of abundance profiles across samples. Color scale indicates log-scaled relative abundances.

**Table 1.**

| Dataset | Citation | Samples<br>(#) | Participants<br>(#) | Mean Reads per<br>Sample |
| --- | --- | --- | --- | --- |
| Kublin | This study | 94 | 15 | 61,722,891.79 |
| Bender-PRJNA521878 | <sup>1</sup> | 62 | 29 | 4,166,371.97 |
| Backhed-PRJEB6456 | <sup>2</sup> | 400 | 100 | 39,764,708.24 |
| Nguyen-PRJNA630999 | <sup>3</sup> | 292 | 77 | 64,487,422.86 |
| **Yassour-**<br>**PRJNA475246** | 4 | 169 | 43 | 28,045,378.54 |
| **Lou-PRJNA698986** | 5 | 2,049 | 642 | 26,306,197.79 |
| **Leonard-**<br>**PRJNA486782** | 6 | 96 | 24 | 43,482,751.56 |
| **Eng-PRJNA510445** | 7 | 1,279 | 232 | 27,790,671.30 |

Summary of metagenomic WGS gut microbiome datasets included in meta-analysis.

To focus our analysis on CAGs most likely to represent species-level groupings of genes, we subset our analysis to those CAGs containing between 1,000 and 10,000 genes (n = 592), a range which encompasses most representative bacterial genomes in the NCBI RefSeq database (Fig. 1B). We conceptualized these CAGs as “genomospecies” because they define a group of organisms at the species or strain level based on a high degree of shared genomic content[61]. The filtered set of 592 appropriately sized CAGs, i.e., genomospecies, account for over half of the raw sequence fragments recovered from infant stool samples with no clear bias by study or timepoint (Fig. 1C). While the organisms contributing the remaining sequence fragments may also have a meaningful influence on human health, they were not observed consistently at high enough abundance across multiple samples to enable gene-level analysis in this study. Using these genomospecies-level abundances as the basis of characterizing microbiome composition in our to -cohort, we compared pairs of samples from the same or different individuals using rank correlations across the CAGs. Sample pairs from the same individual were more correlated (Fig. 1D, p=1.16E-46 Mann-Whitney U), as were samples collected from the same individual at shorter time intervals compared to longer intervals (Fig. 1E, p=4.12E-10 Spearman); together these show that there is some degree of temporal stability in community composition. A graphical summary of microbial abundances across samples is shown in Figure 1F, with each genomospecies shown in a row and each sample shown in a column. Samples are ordered by participant and timepoint, and the genomospecies are grouped by linkage clustering based on the similarity of abundance patterns across samples. The presence of genomospecies with very similar patterns of abundance in this dataset suggests that organisms are not distributed randomly across individuals, but that there may be groups of genomospecies whose relative abundance are correlated when comparing across specimens (Fig. 1F).

### Bacterial strains are observed in tightly correlated consortia across populations

To identify microbial species with correlated abundances, we calculated the Kendall rank correlation coefficient[62] for every pair of genomospecies across all of the samples from both published and newly-generated microbiome samples. Ordination of genomospecies based on similarity of abundance profiles across samples suggested a correlation within taxonomic groups (Fig. 2A; Supp. Data 1). While correlation coefficients were higher overall within taxonomic groups than between taxonomic groups (ANOSIM R=0.43 p=0.001) this was not observed universally across taxonomic groups, with many CAG pairs from the same taxa showing a complete lack of correlation (Fig. 2B, S1). Having observed that taxonomic similarity was not the primary driver of correlated abundances, we compared all pairs of genomospecies in a taxonomically-agnostic analysis. Out of all pairwise comparisons of genomospecies (n=174,936) only 143 had a Kendall’s tau value of 0.7 or greater (shown with connecting lines in Fig. 2A). The correlation of this subset of CAG pairs remained strong even after filtering the data to only include a single sample from each participant or calculating the correlation independently for each study (Fig. 2C), and so it is not likely to be driven by the confounding effect of intra-individual or inter-population differences in community composition.

**Figure 2.**
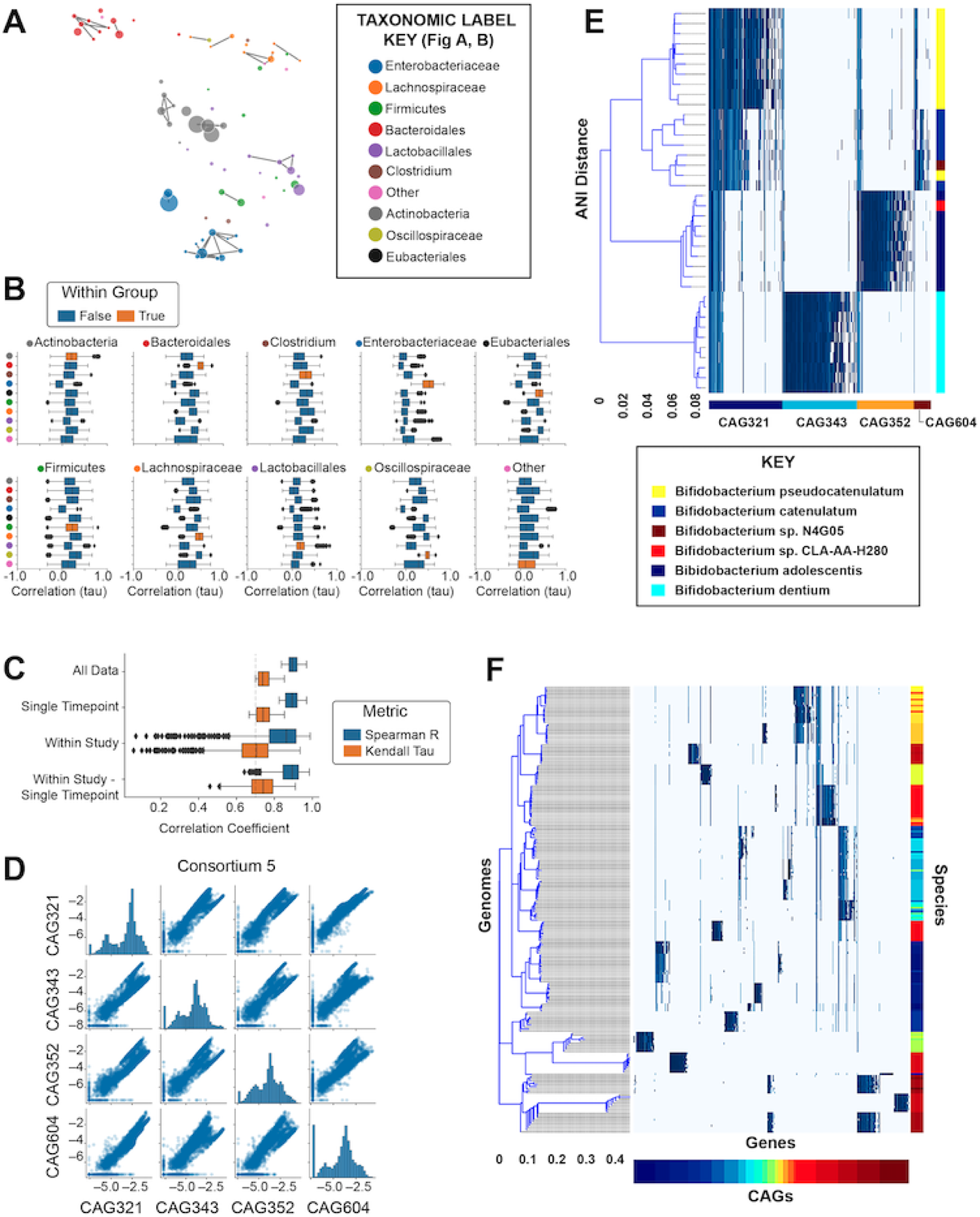
Groups of microbial genomospecies are reproducibly observed at correlated abundances across studies. A) UMAP-based ordination of genomospecies (filtered at a minimum threshold of 0.1% average abundance) based on correlation of relative abundances across samples (Kendall’s tau). B) Comparison of relative abundance-based correlation coefficients for genomospecies pairs based on order-level taxonomic annotations. C) Considering only those pairs of genomospecies with a correlation coefficient greater than 0.7 (Kendall’s tau) using all available data, correlation metrics were recalculated using a single timepoint per participant across all studies (“Single Timepoint”); while calculating an independent correlation metric for each individual study (“Within Study”); or using a single timepoint per participant while also calculating an independent correlation metric for each individual study (“Within Study – Single Timepoint”). The Spearman correlation coefficient was also calculated for all comparisons (blue) in addition to Kendall’s tau (orange). D) Comparison of relative abundances (CLR) across all samples for each pair of genomospecies within Consortium 5. E) Bacterial reference genome similarity for each of the genes within the 4 genomospecies which make up Consortium 5. Each column represents a single gene reconstructed from the metagenomic analysis. The bottom color bar indicates the genomospecies (CAG) assignment for each gene. Blue marks indicate reference genomes (each shown in a distinct row) in which that gene was detected by sequence alignment. The right-hand color bar indicates the species-level assignment for each reference genome. Hierarchical clustering of reference genomes is based on the average nucleotide identity-based dissimilarity matrix. F) Bacterial reference genome similarity for Consortium 3 (following E), with the full set of reference genomes available for inspection in Supplementary Data 2.

Next, groups of microbes were identified by single-linkage clustering using these highly correlated genomospecies, which we conceptualized as “consortia” because of their high degree of co-abundance in the infant gut microbiome (Table S3). We retained all genomospecies in this analysis, with those that did not have any correlated match being treated as individual single-genomospecies “consortia.” The most abundant consortia accounted for 1-5% of all predicted genome copies on average across all specimens in the meta-analysis (Table S4, S5). To test our hypothesis that these highly correlated genomospecies represent multiple organisms (in comparison to the null hypothesis that that a single organism encodes all of the observed co-abundant genes), we compared these metagenome-derived genomospecies to the reference genomes of bacterial isolates. To identify the bacterial reference genomes which are most similar to each genomospecies we searched the NCBI RefSeq collection of bacterial genomes (n=113,938; downloaded June 6^th^, 2022) by amino acid sequence alignment. To better understand genomospecies relationships to conventional phylogenetic based metagenome interpretation, we closely examined the two largest groups of genomospecies with highly correlated abundances (Fig. 2D, S2). We made two observations.

First, the genes contained within each individual genomospecies generally mapped to a consistent set of genomes. Second, genomospecies within those consortia often mapped to different strains and species within a genus (Fig. 2A,2E) or even different orders within a class (Fig. 2F, Supp. Data 2). Finding no single genome with the complete genetic content present in these correlated genomospecies, it is likely that they represent groups of distinct organisms that are present at correlated relative abundances in the human gut microbiome during early life.

### Relative abundance of microbial consortia changes rapidly during human infancy

Considering the human gut microbiome as a collection of microbial consortia, we wanted to better understand how this complex community evolves during early life. Individual consortia vary widely in relative abundance both as a function of host age as well as study population (Fig. 3A-B, Supp. Data 3). Because each study included samples from a single population, it was not possible to distinguish between study population differences and batch effects of sampling. Ordinating samples based on consortium abundances across all studies shows a complex pattern, suggesting that samples at earlier timepoints are more varied in community composition, and samples at later timepoints converge on a smaller number of community types (Fig. 3C, Supp. Data 4). Consistent with this hypothesis, we found that the composition of microbial consortia was more similar at premature-birth and later timepoints within each study (Fig. 3D, p=0.008 Spearman).

**Figure 3.**
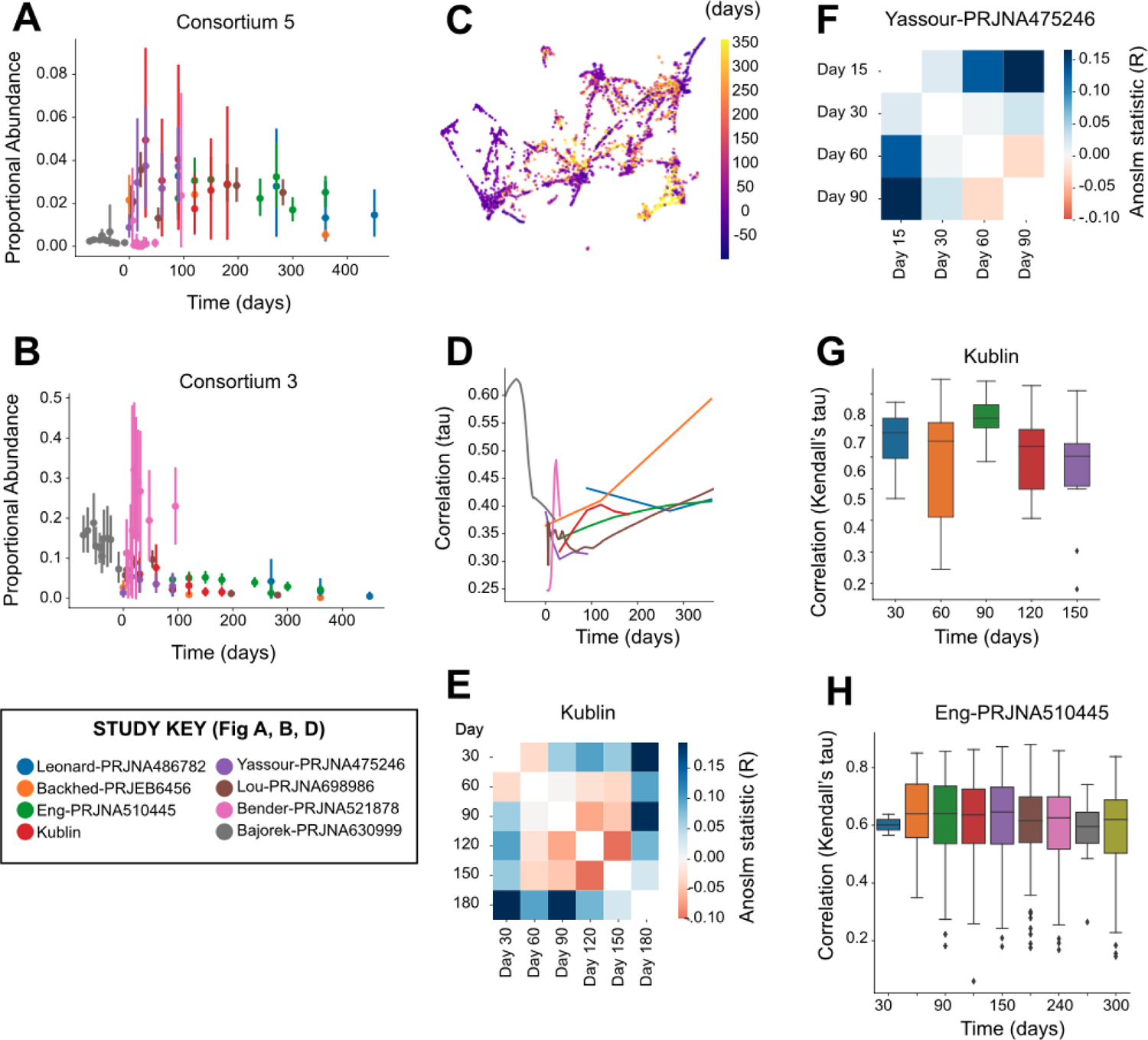
Rapid changes in relative abundance of microbial consortia during early human life. A) Summary of the relative abundance of Consortium 5 (vertical axis) across stool samples as a function of time since term birth (horizontal axis), comparing samples obtained from different studies (indicated by color). B) Summary of the relative abundance for Consortium 3, as in (A). C) UMAP-based ordination of microbiome samples based on similarity of microbiome composition as measured by the relative abundance of microbial consortia. Colors indicate the timepoint of sample collection relative to term birth. D) Similarity of sample composition was compared for pairs of samples collected at similar timepoints from different individuals within each study using Kendall’s tau. The horizontal axis indicates the time of sampling, and the vertical axis indicates the similarity of microbial abundances observed between different individuals. E) Similarity of microbial relative abundances were compared between pairs of samples collected at different timepoints from different individuals within the samples collected for this study (Kublin). The pairwise comparison of each timepoint using the ANOSIM R metric is shown in a heatmap, with positive values indicating more distinct microbial compositions within each of the pair of timepoints and negative values indicating more similar microbial compositions within the pair of timepoints. F) Similarity of microbial relative abundances for the samples from the Yassour study (as in E). G) Similarity of samples collected from the same individual at adjacent timepoints within the samples collected for this study (Kublin). The horizontal axis indicates the timepoint which was compared to samples from the immediately proceeding timepoint. Higher values on the vertical axis indicate a greater similarity of samples based on the relative abundance of microbial consortia. H) Similarity of samples collected from the same individual from the Eng study (as in G).

To better understand the dynamics of microbial communities as a function of host age, we used ANOSIM to compare all the samples collected at each pair of timepoints within each study. In our data, we noted a similarity of days 30-60 as well as days 90-150, with day 180 as the most distinct (Fig. 3E). In contrast, the Yassour et al. dataset showed a greater similarity of days 60-90 than 30-60 (Fig. 3F). Looking entirely at the similarity of samples from the same individual over time, our data showed a greater degree of change (lower correlation) from days 60-90 than 30-60 or 90-120 (Fig. 3G), while the Eng et al. data showed a greater degree of change from 30-60 than 60-90 or 90-120 (Fig. 3H). The combined analysis across cohorts emphasizes the high degree of interpersonal heterogeneity and temporal transience in the human gut microbiome during early life.

### Taxonomically diverse microbial consortia are less abundant in the gut of infants with cystic fibrosis

An important application of detailed microbiome analysis is to identify microbes which may influence human health and disease. A study published by Eng, et al.[53] paired metagenomic sequencing with infant health indicators and compared the metabolic pathways encoded by the microbiome with inflammation and nutritional failure in We accessed a rich dataset that paired data from infants with cystic fibrosis (CF; n=207) to healthy controls (n=25). As previously observed[63], infants with CF had lower weight than healthy controls at each timepoint (Fig. 4A, Wilcoxon p=0.0017). To identify organisms with relative abundances that are correlated with CF status and/or weight, we performed independent linear modeling at each timepoint. The weight-association analysis was performed using only samples from participants with CF. Because of uneven sampling between groups, the CF- and weight-association analyses were performed over an overlapping but distinct set of days. When comparing the strength of association with these two clinical features across the consortia, we observed that the organisms with positive weight-associations (observed at higher abundances in CF infants with greater weights) generally had negative CF-associations (observed at lower abundances in CF infants compared to healthy controls), and vice versa (Fig. 4B-C).

**Figure 4.**
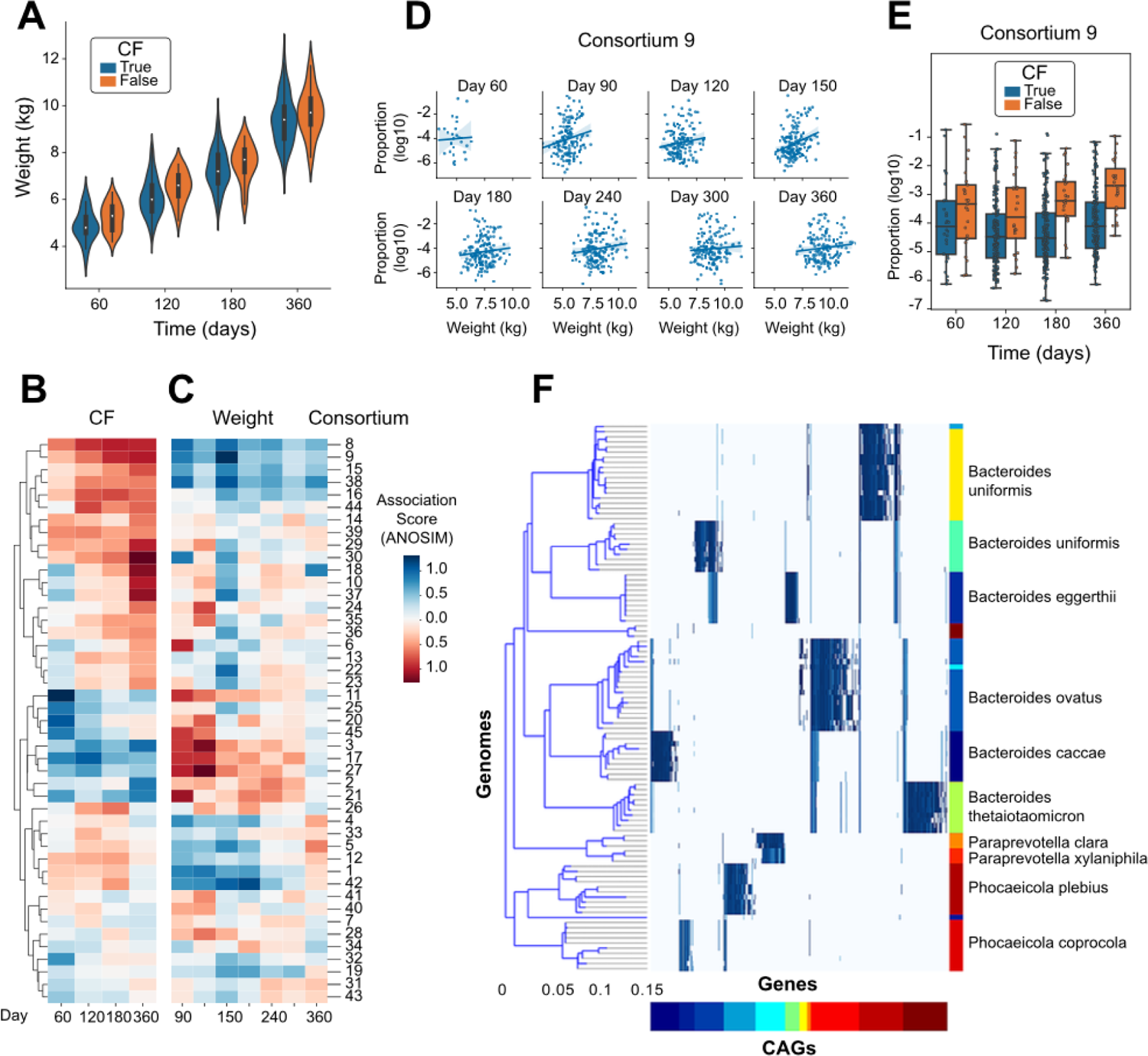
Association of specific microbial consortia with infant CF status and weight. A) Measured weight of each infant at each timepoint, distinguishing infants diagnosed with CF from healthy controls. B) Estimated coefficient of association for the relative abundance of each microbial consortium with CF status, calculated independently at each timepoint. C) Estimated coefficient of association for the relative abundance of each microbial consortium with infant weight using only those participants diagnosed with CF, calculated independently at each timepoint. D) Comparison of the relative abundance of Consortium 9 with weight within the group of participants diagnosed with CF, shown independently at each timepoint. E) Comparison of the relative abundance of Consortium 9 between participants distinguished by CF diagnosis, shown independently at each timepoint. F) Comparison of the genomic content of Consortium 9 to a reference genome collection, as in Figure 2E.

The microbial consortium showing the strongest association with weight in the CF infants was #9 (Fig. 4D), which was also found at lower abundance in CF infants compared to healthy controls (Fig. 4E). Alignment of the genomic markers of consortium #9 against the NCBI RefSeq catalog of microbial genomes identified species spanning *Bacteroides* (*B. uniformis*, *B. stercoris*, *B. eggerthii*, *B. ovatus*, *B. caccae*, and *B. thetaiotaomicron*), *Paraprevotella clara/xylaniphila*, and *Phocaeicola* (*P. plebius* and *P. coprocola*) (Fig 4F). While these genera have been identified previously as being altered in the gut microbiome of infants with CF, these resultsi: (a) identify the species that are most likely involved with a specific set of genomic markers (Supp. Data 5), (ii) indicate that those species are generally found together rather than individually in the gut microbiome, and (iii) suggest that the combined metabolism of a multi-species consortium may collectively mediate weight gain in infants with CF.

## Discussion

### Quantification of microbes sampled from the human gut

By using the reference-free analysis approach to microbiome analysis implemented in the *geneshot* analysis pipeline[60], our analysis aimed to expand our understanding of the infant gut microbiome. The advantage of this approach, which quantifies organisms on the basis of the genes encoded in their genome, is that it is not dependent on the composition of existing genome databases to detect and quantify specific organisms. While the primary drawback of this approach is a lack of sensitivity for the detection of organisms that are not sequenced to a depth sufficient for *de novo* reconstruction, approximately 50% of the raw metagenomic data was successfully assigned to just 592 distinct genomospecies representing the most abundant organisms (Fig. 1C). Based on previous work, we expected that microbial composition would reflect some degree of individuality and temporal stability[55, 64, 65]. This expectation was borne out using the genomospecies-level abundance data, with samples more similar within-than between-participants (Fig. 1D) and more similar between samples collected across shorter time intervals (Fig. 1E). Based on these high-level metrics, we gained confidence that our genomospecies-level analysis is capturing a biologically meaningful profile of the most abundant organisms in the infant gut microbiome.

### Observation of taxonomically distinct microbial consortia

In addition to the detection of previously unsequenced organisms, an advantage of *de novo* metagenomic analysis is the ability to precisely identify organisms with correlated abundances that are taxonomically similar or diverse. While marker-gene or *k*-mer based analyses run the risk of confounding taxa that share a subset of genomic content, our *de novo* gene-level analysis assigns each raw sequence read unambiguously to a single genomospecies reference (using an expectation maximization approach to resolve duplicate alignments). Moreover, by limiting to the 592 organisms, evaluating all possible pairwise correlations among microbes became computationally tractable. Using this approach, we found only 143 pairs of microbes (out of the 174,936 total pairwise comparisons) with a Kendall’s tau correlation coefficient ≥0.7. Noting the inter-participant individuality of microbiome composition (Fig. 1D), we were encouraged that this high degree of pairwise correlation between individual genomospecies was observed after downsampling to a single sample per participant and after controlling for batch effects (Fig. 1C).

While it is possible that genomospecies with correlated abundances may represent a single species which was inappropriately split due to noise in the metagenomic sequencing process, it is more likely that correlated genomospecies represent different species with correlated abundances. One biological concept used to describe such multi-species groups would be “consortia” of distinct organisms formed by cross-feeding or syntrophic interaction[66] or by stable niche partitioning of a common source of energy (such as the degradation of diverse human milk oligosaccharides by related *Bifidobacteria*[67]). By comparing each genomospecies’ genetic content to the extensive NCBI RefSeq genome collection, we observed candidate consortia containing genetically distinct organisms from the same genus (e.g., Consortium 5, Fig. 2E, Supp. Data 2), as well as single consortia containing organisms spanning multiple diverse genera (*Klebsiella, Enterobacter, Leclercia, Citrobacter, Cronobacter, Proteus, Serratia,* and *Pseudomonas*) (e,g, Consortium 3, Fig. 2F, Supp. Data 2). The robust correlation of relative abundances between these genetically distinct organisms is highly unlikely to be caused by technical artifacts, and strongly suggests that these groups of organisms are present or absent in the microbiome as a correlated group.

### Complex, rapid temporal dynamics of the infant gut microbiome

Using the aggregate abundances of microbial consortia to measure the composition of the gut microbiome, we sought to better understand the complex temporal dynamics of the developing human microbiome during infancy. While there were some consistent patterns across datasets – consortium #5 of *Bifidobacteria* was observed at higher abundance during later timepoints (Fig. 3A) and consortium #3 of diverse *Enterobacterales* observed at higher abundance during earlier timepoints (Fig. 3B) – those patterns were not consistent across all individuals or all studies. Clustering of samples by total community composition did not reveal any single community state associated with earlier or later timepoints (Fig. 3C). The most consistent pattern we observed was that microbial communities from later timepoints were more similar across individuals than the communities from later timepoints, an effect which was observed across multiple independent studies (Fig. 3D) and which is consistent with the previous observations made within single cohorts [11, 54, 55].

The development of the human microbiome during the earliest days of life is a highly dynamic process which has not been measured at consistent, dense intervals across previous studies. We augmented the published set of microbiome studies by collecting stool samples at 30-day intervals from birth to day 180 in a cohort of 15 infants. While the stool microbiome in this cohort was more similar between days 30-60 and 90-150 (Fig. 3E), a previous study has shown a stronger signal of similarity between days 60-90 (Fig. 3F). To identify the patterns of microbiome development that are consistent across populations, the field will need to collect considerably more metagenomic data at higher temporal frequency from this early time period across multiple geographically diverse study sites.

### Identification of a multispecies microbial consortia associated with health outcomes in infants with cystic fibrosis

To assess whether any of the newly-identified microbial consortia were correlated with human health outcomes, we focused on a study of infants with cystic fibrosis [53], performing linear modeling of consortium abundances with both CF status and weight among infants with CF (those analyses being performed independently). The strongest association was observed with a multispecies consortium (#9) containing species of *Bacteroides*, *Paraprevotella*, and *Phocaeicola* (Fig. 4F). This group of organisms was found at lower abundance in infants with CF compared to healthy controls; and critically was also found at lower abundance in those infants with CF who weighed less; in particular at day 90 of life (Fig. 4B-E). Our biological interpretation, which is heavily influenced by the identification of this microbial consortium, is that there is a mechanistic link between the presence of this group of microbes with the factors influencing weight gain during early life. Similar to the joint metabolism of human milk oligosaccharides distributed across related *Bifidobacteria*[67], we hypothesize that there is a consequence from the presence of this group of organisms which may not be recapitulated by any single member. When translating these findings to a controlled experimental setting, our results would imply that the administration of any single species may not be sufficient to reproduce the same biological effect, but instead the full or partial set of the multi-species community may be required.

## Methods

### Study sites and enrollment of cohort

We obtained data from a collaborative mother-infant cohort that enrolled pregnant women in the Guangdong and Zhejiang Provinces of China from and followed their newborn off-spring from birth up to two years of age[56, 68]. Samples were collected for this analysis from 12/19/2017 to 08/21/2018. Pregnant women provided written informed consent and were screened and enrolled between 14 and 20 weeks of gestation. The study completed enrollment into the cohort in January, 2020.

### Specimen and data collection and processing

Stool samples were initially collected at the hospital (10 grams of fecal matter collected from diapers, placed in plastic containers and stored at −80°C). Subsequent stool samples were collected monthly by parents in the home. On the morning of sample collection, a cooler with ice packs and sample collection materials was delivered to the home of each participant. In the early evening, the coolers were collected and returned to the laboratory where samples were aliquoted, labelled and stored at −80°C. If an infant did not provide a stool sample on the collection day, collection was rescheduled for the following day and a new cooler was provided.

### Sequencing

DNA was extracted from each stool sample (n=94) and prepared for sequencing using the TruPrep DNA Library Prep Kit V2 for Illumina. Libraries were clustered and sequenced on an Illumina HiSeq2000 instrument and sequenced to an average depth of 61.7 million paired-end reads per sample.

Sequencing data from published datasets were obtained using the SRA Toolkit, downloading all paired-end FASTQ data available from the BioProject accessions PRJNA521878 (Bender), PRJEB6456 (Backhed), PRJNA630999 (Bajorek), PRJNA475246 (Yassour), PRJNA698986 (Lou), PRJNA486782 (Leonard), and PRJNA510445 (Eng).

## Analysis

### Identifying and quantifying CAGs from metagenomes

While previous reports have described bacteria in broad taxonomic groups (e.g. Bifidobacteriaceae, Lactobacillus, Enterococcus, Bacteroides, Streptococcus), we used gene-level metagenomics to increase this level of resolution to the species- and strain-level, while also identifying horizontally transferred genetic elements which play a role in microbiome development. Analysis of raw FASTQ datasets was performed using the geneshot analysis pipeline, available at https://github.com/Golob-Minot/geneshot. The exact version of that software used was v0.9 with the commit hash 4d700993660ed8fdf4df6432d2c7cb2ddd8ce85f. The geneshot pipeline (described previously [60]) performs the following bioinformatics analysis steps:

1. De novo assembly of each sample independently (using megahit v1.2.9);
2. Identification of protein-coding sequences in each assembly (using prodigal v2.6.3);
3. Deduplication of protein-coding sequences at 90% sequencing identity and 50% coverage (using linclust/MMseqs2 release 12-113e3);
4. Alignment of conceptually translated sequence reads against that deduplicated gene catalog (using DIAMOND v0.9.10);
5. Clustering of protein-coding sequences into CAGs using a maximum cosine distance threshold of 0.35.

To effectively process the large number of metagenomic samples in this project, a subset was selected for *de novo* assembly and gene identification which included only a single representative per participant across all projects, while genes and CAGs were quantified across the full set of samples. The computational resources required for this analysis were considerable, with ∼104,000 CPU hours required for the *de novo* assembly and gene identification and ∼364,000 CPU hours required to align the full dataset against that gene catalog.

### Comparing the composition of microbial communities

The similarity of organisms present in different microbial communities was estimated using the non-parametric Spearman correlation of CLR-transformed abundances. The Spearman R value was used when comparing pairs of samples in terms of their microbial composition. When comparing pairs of CAGs to find organisms with correlated abundances, the more conservative Kendall Tau metric was also calculated using the CLR-transformed abundances.

### Identifying genomospecies associated with health status

Statistical analysis for the association of genomospecies relative abundance with the health status of human hosts (CF status and weight) was performed using Generalized Estimating Equations as implemented in the statsmodels package (Python). All GEE models were constructed using an exchangeable covariance structure and Gaussian family. Adjustment for multiple hypothesis testing was performing with the FDR-BH protocol as implemented in statsmodels.

### Comparing genetic content of genomospecies to reference genomes

The reference genomes most closely resembling the organisms reconstructed *de novo* from this metagenomic dataset were identified by alignment against the NCBI RefSeq database (downloaded June 6, 2022). The protein-coding sequences from each CAG (“genomospecies”) was aligned against that genome collection using the gig-map workflow (available at https://github.com/FredHutch/gig-map/), which employs the DIAMOND aligner for rapid alignment of conceptually-translated genomes in amino acid space, at a minimum alignment threshold of 90% sequence identity and 90% alignment coverage (of the *de novo* assembled gene sequence). The similarity of reference genomes (used for the dendrogram display in CAG-genome heatmaps) was estimated by gig-map with Average Nucleotide Identity (ANI) calculated using the MASH software [69].

## Data Availability

The data underlying the results presented in the study are available from the Cirro Data Platform (https://cirro.bio) using anonymous login information – username: password: Public_Manuscript_Data123.

## Supplementary Tables

- Supplementary Table 1: Manifest with metadata for all specimens, including number of reads, number of genes detected, etc.
- Supplementary Table 2: Relative abundance of all genomes across all samples
- Supplementary Table 3: Annotation of genomes by consortium
- Supplementary Table 4: Annotation of consortia, mean relative abundance
- Supplementary Table 5: Relative abundance of all consortia across all samples

## Supplementary Figures

- Supplementary Figure 1: Kendall’s tau within the most frequent taxonomic groups at various levels

## Supplementary Data

- Supplementary Data 1: Interactive display showing ordination of CAGs with taxonomic annotations
- Supplementary Data 2: gig-map displays for all consortia
- Supplementary Data 3: Relative abundance displays over time and across studies for all consortia
- Supplementary Data 4: Interactive UMAP of samples by consortium abundance
- Supplementary Data 5: Sequences of genes in all CAGs

## Declarations

### Ethics approval and consent to participate

The study was approved by the ethical committee of the Chinese Center for Disease Control and Prevention.

### Consent for publication

Not applicable

### Availability of data and materials

The data produced by this study has been made available in the Cirro Data Platform (https://cirro.bio) using anonymous login information – username; password: Public_Manuscript_Data123. To ensure the privacy of all participants, all sequences from the metagenomic data which align to the human genome have been masked from the FASTQ files which are provided publicly. Note that Supplementary Data 5 is only available via the Cirro platform due to the large file size (870MB). The datasets supporting the conclusions of this article are included within the article and its additional files.

### Competing interests

The authors declare that they have no competing interests.

## Funding

SM was supported by funding from the Microbiome Research Initiative (Fred Hutch Cancer Center, PI: David Fredricks M.D.). SM, KMB, AFG, AJ, and JK were supported by funding from NIH NIAID R01AI127100.

## Authors’ contributions

JK, XS, PB, and SS conceived and designed the study for the newly collected cohort. XS, LL, and YJ analyzed and generated data from physical specimens. SM performed bioinformatics analysis and implemented statistical analyses. SM, KMB, AFG, AJ, and JK collaboratively developed the statistical analysis approach. SM, PB, KMB, AFG, and JK collaboratively wrote the manuscript. All authors read and approved the final manuscript.

## Acknowledgements

We would like to acknowledge the significant contribution of all study participants.

## Notes

### Competing Interest Statement

The authors have declared no competing interest.

### Funding Statement

Yes

### Author Declarations

Medical ethical review committee, national institute for occupational health and poison control, chinese center for disease control and prevention, approval number is 201718. Written informed consent was obtained at time of enrollment.

## References

1. Wang S, Ryan CA, Boyaval P, Dempsey EM, Ross RP, Stanton C: Maternal Vertical Transmission Affecting Early-life Microbiota Development. Trends Microbiol 2020, 28:28–45.

2. Hayden HS, Eng A, Pope CE, Brittnacher MJ, Vo AT, Weiss EJ, Hager KR, Martin BD, Leung DH, Heltshe SL, et al: Fecal dysbiosis in infants with cystic fibrosis is associated with early linear growth failure. Nat Med 2020, 26:215–221.

3. Ennamorati M, Vasudevan C, Clerkin K, Halvorsen S, Verma S, Ibrahim S, Prosper S, Porter C, Yeliseyev V, Kim M, et al: Intestinal microbes influence development of thymic lymphocytes in early life. Proc Natl Acad Sci U S A 2020, 117:2570–2578.

4. Backhed F, Roswall J, Peng Y, Feng Q, Jia H, Kovatcheva-Datchary P, Li Y, Xia Y, Xie H, Zhong H, et al: Dynamics and Stabilization of the Human Gut Microbiome during the First Year of Life. Cell Host Microbe 2015, 17:690–703.

5. Turnbaugh PJ, Hamady M, Yatsunenko T, Cantarel BL, Duncan A, Ley RE, Sogin ML, Jones WJ, Roe BA, Affourtit JP, et al: A core gut microbiome in obese and lean twins. Nature 2009, 457:480–484.

6. Yassour M, Jason E, Hogstrom LJ, Arthur TD, Tripathi S, Siljander H, Selvenius J, Oikarinen S, Hyoty H, Virtanen SM, et al: Strain-Level Analysis of Mother-to-Child Bacterial Transmission during the First Few Months of Life. Cell Host Microbe 2018, 24:146–154 e144.

7. Edwards KM: Maternal antibodies and infant immune responses to vaccines. Vaccine 2015, 33:6469–6472.

8. Voysey M, Kelly DF, Fanshawe TR, Sadarangani M, O’Brien KL, Perera R, Pollard AJ: The Influence of Maternally Derived Antibody and Infant Age at Vaccination on Infant Vaccine Responses: An Individual Participant Meta-analysis. JAMA Pediatr 2017, 171:637–646.

9. Zimmermann P, Curtis N: Factors That Influence the Immune Response to Vaccination. Clin Microbiol Rev 2019, 32.

10. Tsang JS, Dobano C, VanDamme P, Moncunill G, Marchant A, Othman RB, Sadarangani M, Koff WC, Kollmann TR: Improving Vaccine-Induced Immunity: Can Baseline Predict Outcome? Trends Immunol 2020, 41:457–465.

11. Kostic AD, Gevers D, Siljander H, Vatanen T, Hyotylainen T, Hamalainen AM, Peet A, Tillmann V, Poho P, Mattila I, et al: The dynamics of the human infant gut microbiome in development and in progression toward type 1 diabetes. Cell Host Microbe 2015, 17:260–273.

12. Gray J, Oehrle K, Worthen G, Alenghat T, Whitsett J, Deshmukh H: Intestinal commensal bacteria mediate lung mucosal immunity and promote resistance of newborn mice to infection. Sci Transl Med 2017, 9.

13. Kim YG, Sakamoto K, Seo SU, Pickard JM, Gillilland MG, 3rd, Pudlo NA, Hoostal M, Li X, Wang TD, Feehley T, et al: Neonatal acquisition of Clostridia species protects against colonization by bacterial pathogens. Science 2017, 356:315–319.

14. Fujimura KE, Sitarik AR, Havstad S, Lin DL, Levan S, Fadrosh D, Panzer AR, LaMere B, Rackaityte E, Lukacs NW, et al: Neonatal gut microbiota associates with childhood multisensitized atopy and T cell differentiation. Nat Med 2016, 22:1187–1191.

15. Vatanen T, Franzosa EA, Schwager R, Tripathi S, Arthur TD, Vehik K, Lernmark A, Hagopian WA, Rewers MJ, She JX, et al: The human gut microbiome in early-onset type 1 diabetes from the TEDDY study. Nature 2018, 562:589–594.

16. Vatanen T, Kostic AD, d’Hennezel E, Siljander H, Franzosa EA, Yassour M, Kolde R, Vlamakis H, Arthur TD, Hamalainen AM, et al: Variation in Microbiome LPS Immunogenicity Contributes to Autoimmunity in Humans. Cell 2016, 165:842–853.

17. Arrieta MC, Stiemsma LT, Dimitriu PA, Thorson L, Russell S, Yurist-Doutsch S, Kuzeljevic B, Gold MJ, Britton HM, Lefebvre DL, et al: Early infancy microbial and metabolic alterations affect risk of childhood asthma. Sci Transl Med 2015, 7:307ra152.

18. Abdel-Gadir A, Stephen-Victor E, Gerber GK, Noval Rivas M, Wang S, Harb H, Wang L, Li N, Crestani E, Spielman S, et al: Microbiota therapy acts via a regulatory T cell MyD88/RORgammat pathway to suppress food allergy. Nat Med 2019, 25:1164–1174.

19. Cahenzli J, Koller Y, Wyss M, Geuking MB, McCoy KD: Intestinal microbial diversity during early-life colonization shapes long-term IgE levels. Cell Host Microbe 2013, 14:559–570.

20. Brand S, Teich R, Dicke T, Harb H, Yildirim AO, Tost J, Schneider-Stock R, Waterland RA, Bauer UM, von Mutius E, et al: Epigenetic regulation in murine offspring as a novel mechanism for transmaternal asthma protection induced by microbes. J Allergy Clin Immunol 2011, 128:618–625 e611-617.

21. Herbst T, Sichelstiel A, Schar C, Yadava K, Burki K, Cahenzli J, McCoy K, Marsland BJ, Harris NL: Dysregulation of allergic airway inflammation in the absence of microbial colonization. Am J Respir Crit Care Med 2011, 184:198–205.

22. Olszak T, An D, Zeissig S, Vera MP, Richter J, Franke A, Glickman JN, Siebert R, Baron RM, Kasper DL, Blumberg RS: Microbial exposure during early life has persistent effects on natural killer T cell function. Science 2012, 336:489–493.

23. Trompette A, Gollwitzer ES, Yadava K, Sichelstiel AK, Sprenger N, Ngom-Bru C, Blanchard C, Junt T, Nicod LP, Harris NL, Marsland BJ: Gut microbiota metabolism of dietary fiber influences allergic airway disease and hematopoiesis. Nat Med 2014, 20:159–166.

24. Deshmukh HS, Liu Y, Menkiti OR, Mei J, Dai N, O’Leary CE, Oliver PM, Kolls JK, Weiser JN, Worthen GS: The microbiota regulates neutrophil homeostasis and host resistance to Escherichia coli K1 sepsis in neonatal mice. Nat Med 2014, 20:524–530.

25. Constantinides MG, Link VM, Tamoutounour S, Wong AC, Perez-Chaparro PJ, Han SJ, Chen YE, Li K, Farhat S, Weckel A, et al: MAIT cells are imprinted by the microbiota in early life and promote tissue repair. Science 2019, 366.

26. Al Nabhani Z, Dulauroy S, Marques R, Cousu C, Al Bounny S, Dejardin F, Sparwasser T, Berard M, Cerf-Bensussan N, Eberl G: A Weaning Reaction to Microbiota Is Required for Resistance to Immunopathologies in the Adult. Immunity 2019, 50:1276–1288 e1275.

27. Pronovost GN, Hsiao EY: Perinatal Interactions between the Microbiome, Immunity, and Neurodevelopment. Immunity 2019, 50:18–36.

28. Knoop KA, Gustafsson JK, McDonald KG, Kulkarni DH, Coughlin PE, McCrate S, Kim D, Hsieh CS, Hogan SP, Elson CO, et al: Microbial antigen encounter during a preweaning interval is critical for tolerance to gut bacteria. Sci Immunol 2017, 2.

29. Scanlan PD, Buckling A, Kong W, Wild Y, Lynch SV, Harrison F: Gut dysbiosis in cystic fibrosis. J Cyst Fibros 2012, 11:454–455.

30. Nielsen S, Needham B, Leach ST, Day AS, Jaffe A, Thomas T, Ooi CY: Disrupted progression of the intestinal microbiota with age in children with cystic fibrosis. Sci Rep 2016, 6:24857.

31. Manor O, Levy R, Pope CE, Hayden HS, Brittnacher MJ, Carr R, Radey MC, Hager KR, Heltshe SL, Ramsey BW, et al: Metagenomic evidence for taxonomic dysbiosis and functional imbalance in the gastrointestinal tracts of children with cystic fibrosis. Sci Rep 2016, 6:22493.

32. Madan JC, Koestler DC, Stanton BA, Davidson L, Moulton LA, Housman ML, Moore JH, Guill MF, Morrison HG, Sogin ML, et al: Serial analysis of the gut and respiratory microbiome in cystic fibrosis in infancy: interaction between intestinal and respiratory tracts and impact of nutritional exposures. mBio 2012, 3.

33. Hoffman LR, Pope CE, Hayden HS, Heltshe S, Levy R, McNamara S, Jacobs MA, Rohmer L, Radey M, Ramsey BW, et al: Escherichia coli dysbiosis correlates with gastrointestinal dysfunction in children with cystic fibrosis. Clin Infect Dis 2014, 58:396–399.

34. Hoen AG, Li J, Moulton LA, O’Toole GA, Housman ML, Koestler DC, Guill MF, Moore JH, Hibberd PL, Morrison HG, et al: Associations between Gut Microbial Colonization in Early Life and Respiratory Outcomes in Cystic Fibrosis. J Pediatr 2015, 167:138–147 e131-133.

35. Vernocchi P, Del Chierico F, Russo A, Majo F, Rossitto M, Valerio M, Casadei L, La Storia A, De Filippis F, Rizzo C, et al: Gut microbiota signatures in cystic fibrosis: Loss of host CFTR function drives the microbiota enterophenotype. PLoS One 2018, 13:e0208171.

36. Duytschaever G, Huys G, Bekaert M, Boulanger L, De Boeck K, Vandamme P: Cross-sectional and longitudinal comparisons of the predominant fecal microbiota compositions of a group of pediatric patients with cystic fibrosis and their healthy siblings. Appl Environ Microbiol 2011, 77:8015–8024.

37. Duytschaever G, Huys G, Bekaert M, Boulanger L, De Boeck K, Vandamme P: Dysbiosis of bifidobacteria and Clostridium cluster XIVa in the cystic fibrosis fecal microbiota. J Cyst Fibros 2013, 12:206–215.

38. Schippa S, Iebba V, Santangelo F, Gagliardi A, De Biase RV, Stamato A, Bertasi S, Lucarelli M, Conte MP, Quattrucci S: Cystic fibrosis transmembrane conductance regulator (CFTR) allelic variants relate to shifts in faecal microbiota of cystic fibrosis patients. PLoS One 2013, 8:e61176.

39. Antosca KM, Chernikova DA, Price CE, Ruoff KL, Li K, Guill MF, Sontag NR, Morrison HG, Hao S, Drumm ML, et al: Altered Stool Microbiota of Infants with Cystic Fibrosis Shows a Reduction in Genera Associated with Immune Programming from Birth. J Bacteriol 2019, 201.

40. Dorsey J, Gonska T: Bacterial overgrowth, dysbiosis, inflammation, and dysmotility in the Cystic Fibrosis intestine. J Cyst Fibros 2017, 16 Suppl 2:S14–S23.

41. Fridge JL, Conrad C, Gerson L, Castillo RO, Cox K: Risk factors for small bowel bacterial overgrowth in cystic fibrosis. J Pediatr Gastroenterol Nutr 2007, 44:212–218.

42. Lisowska A, Madry E, Pogorzelski A, Szydlowski J, Radzikowski A, Walkowiak J: Small intestine bacterial overgrowth does not correspond to intestinal inflammation in cystic fibrosis. Scand J Clin Lab Invest 2010, 70:322–326.

43. Lisowska A, Wojtowicz J, Walkowiak J: Small intestine bacterial overgrowth is frequent in cystic fibrosis: combined hydrogen and methane measurements are required for its detection. Acta Biochim Pol 2009, 56:631–634.

44. Coffey MJ, Nielsen S, Wemheuer B, Kaakoush NO, Garg M, Needham B, Pickford R, Jaffe A, Thomas T, Ooi CY: Gut Microbiota in Children With Cystic Fibrosis: A Taxonomic and Functional Dysbiosis. Sci Rep 2019, 9:18593.

45. Stefka AT, Feehley T, Tripathi P, Qiu J, McCoy K, Mazmanian SK, Tjota MY, Seo GY, Cao S, Theriault BR, et al: Commensal bacteria protect against food allergen sensitization. Proc Natl Acad Sci U S A 2014, 111:13145–13150.

46. Atarashi K, Tanoue T, Oshima K, Suda W, Nagano Y, Nishikawa H, Fukuda S, Saito T, Narushima S, Hase K, et al: Treg induction by a rationally selected mixture of Clostridia strains from the human microbiota. Nature 2013, 500:232–236.

47. Atarashi K, Tanoue T, Shima T, Imaoka A, Kuwahara T, Momose Y, Cheng G, Yamasaki S, Saito T, Ohba Y, et al: Induction of colonic regulatory T cells by indigenous Clostridium species. Science 2011, 331:337–341.

48. Steed AL, Christophi GP, Kaiko GE, Sun L, Goodwin VM, Jain U, Esaulova E, Artyomov MN, Morales DJ, Holtzman MJ, et al: The microbial metabolite desaminotyrosine protects from influenza through type I interferon. Science 2017, 357:498–502.

49. Bender JM, Li F, Purswani H, Capretz T, Cerini C, Zabih S, Hung L, Francis N, Chin S, Pannaraj PS, Aldrovandi G: Early exposure to antibiotics in the neonatal intensive care unit alters the taxonomic and functional infant gut microbiome. J Matern Fetal Neonatal Med 2021, 34:3335–3343.

50. Nguyen M, Holdbrooks H, Mishra P, Abrantes MA, Eskew S, Garma M, Oca CG, McGuckin C, Hein CB, Mitchell RD, et al: Impact of Probiotic B. infantis EVC001 Feeding in Premature Infants on the Gut Microbiome, Nosocomially Acquired Antibiotic Resistance, and Enteric Inflammation. Front Pediatr 2021, 9:618009.

51. Lou YC, Olm MR, Diamond S, Crits-Christoph A, Firek BA, Baker R, Morowitz MJ, Banfield JF: Infant gut strain persistence is associated with maternal origin, phylogeny, and traits including surface adhesion and iron acquisition. Cell Rep Med 2021, 2:100393.

52. Leonard MM, Karathia H, Pujolassos M, Troisi J, Valitutti F, Subramanian P, Camhi S, Kenyon V, Colucci A, Serena G, et al: Multi-omics analysis reveals the influence of genetic and environmental risk factors on developing gut microbiota in infants at risk of celiac disease. Microbiome 2020, 8:130.

53. Eng A, Hayden HS, Pope CE, Brittnacher MJ, Vo AT, Weiss EJ, Hager KR, Leung DH, Heltshe SL, Raftery D, et al: Infants with cystic fibrosis have altered fecal functional capacities with potential clinical and metabolic consequences. BMC Microbiol 2021, 21:247.

54. Stewart CJ, Ajami NJ, O’Brien JL, Hutchinson DS, Smith DP, Wong MC, Ross MC, Lloyd RE, Doddapaneni H, Metcalf GA, et al: Temporal development of the gut microbiome in early childhood from the TEDDY study. Nature 2018, 562:583–588.

55. Koenig JE, Spor A, Scalfone N, Fricker AD, Stombaugh J, Knight R, Angenent LT, Ley RE: Succession of microbial consortia in the developing infant gut microbiome. Proc Natl Acad Sci U S A 2011, 108 Suppl 1:4578–4585.

56. Yao L, Liu L, Dong M, Yang J, Zhao Z, Chen J, Lv L, Wu Z, Wang J, Sun X, et al: Trimester-specific prenatal heavy metal exposures and sex-specific postpartum size and growth. J Expo Sci Environ Epidemiol 2022.

57. Coelho LP, Alves R, Del Rio AR, Myers PN, Cantalapiedra CP, Giner-Lamia J, Schmidt TS, Mende DR, Orakov A, Letunic I, et al: Towards the biogeography of prokaryotic genes. Nature 2022, 601:252–256.

58. Nielsen HB, Almeida M, Juncker AS, Rasmussen S, Li J, Sunagawa S, Plichta DR, Gautier L, Pedersen AG, Le Chatelier E, et al: Identification and assembly of genomes and genetic elements in complex metagenomic samples without using reference genomes. Nat Biotechnol 2014, 32:822–828.

59. Minot SS, Willis AD: Clustering co-abundant genes identifies components of the gut microbiome that are reproducibly associated with colorectal cancer and inflammatory bowel disease. Microbiome 2019, 7:110.

60. Minot SS, Barry KC, Kasman C, Golob JL, Willis AD: geneshot: gene-level metagenomics identifies genome islands associated with immunotherapy response. Genome Biol 2021, 22:135.

61. Brenner DJ, Grimont PA, Steigerwalt AG, Fanning GR, Ageron E, Riddle CF: Classification of citrobacteria by DNA hybridization: designation of Citrobacter farmeri sp. nov., Citrobacter youngae sp. nov., Citrobacter braakii sp. nov., Citrobacter werkmanii sp. nov., Citrobacter sedlakii sp. nov., and three unnamed Citrobacter genomospecies. Int J Syst Bacteriol 1993, 43:645–658.

62. Kendall MG: A New Measure of Rank Correlation. Biometrika 1938, 30:81–93.

63. Patterson KD, Kyriacou T, Desai M, Carroll WD, Gilchrist FJ: Factors affecting the growth of infants diagnosed with cystic fibrosis by newborn screening. BMC Pediatr 2019, 19:356.

64. Yassour M, Vatanen T, Siljander H, Hamalainen AM, Harkonen T, Ryhanen SJ, Franzosa EA, Vlamakis H, Huttenhower C, Gevers D, et al: Natural history of the infant gut microbiome and impact of antibiotic treatment on bacterial strain diversity and stability. Sci Transl Med 2016, 8:343ra381.

65. Vatanen T, Plichta DR, Somani J, Munch PC, Arthur TD, Hall AB, Rudolf S, Oakeley EJ, Ke X, Young RA, et al: Genomic variation and strain-specific functional adaptation in the human gut microbiome during early life. Nat Microbiol 2019, 4:470–479.

66. Ferry JG, Wolfe RS: Anaerobic degradation of benzoate to methane by a microbial consortium. Arch Microbiol 1976, 107:33–40.

67. Lawson MAE, O’Neill IJ, Kujawska M, Gowrinadh Javvadi S, Wijeyesekera A, Flegg Z, Chalklen L, Hall LJ: Breast milk-derived human milk oligosaccharides promote Bifidobacterium interactions within a single ecosystem. ISME J 2020, 14:635–648.

68. Liu L, Yao L, Dong M, Liu T, Lai W, Yin X, Zhou S, Lv L, Li L, Wang J, et al: Maternal urinary cadmium concentrations in early pregnancy in relation to prenatal and postpartum size of offspring. J Trace Elem Med Biol 2021, 68:126823.

69. Ondov BD, Treangen TJ, Melsted P, Mallonee AB, Bergman NH, Koren S, Phillippy AM: Mash: fast genome and metagenome distance estimation using MinHash. Genome Biol 2016, 17:132.

